# Budget Impact Analysis of Disease-Modifying Therapies for Alzheimer’s Disease in Ireland: A Model-Based Analysis

**DOI:** 10.64898/2025.12.30.25343198

**Authors:** Sevinç Elif Şen

## Abstract

**Background:** Disease-modifying therapies (DMTs) for Alzheimer’s disease, including lecanemab and donanemab, have received regulatory approval in multiple jurisdictions. These therapies require complex diagnostic workup and safety monitoring, raising significant budget impact concerns for healthcare payers. No budget impact analysis specific to Ireland or comparable small European healthcare systems has been published.

**Objective:** To estimate the 5-year budget impact of introducing DMTs for early-stage Alzheimer’s disease in Ireland from the Health Service Executive (HSE) payer perspective.

**Methods:** A budget impact model was developed following International Society for Pharmacoeconomics and Outcomes Research (ISPOR) guidelines. The model incorporated Irish epidemiological data, published drug prices, and healthcare resource utilisation estimates. Three treatment uptake scenarios (conservative, base case, optimistic) were modelled over a 5-year time horizon. Sensitivity analyses examined parameter uncertainty.

**Results:** From an estimated eligible population of 11,568 individuals with early-stage Alzheimer’s disease, annual budget impact in Year 5 ranged from €12.8 million (conservative: 3% uptake) to €89.6 million (optimistic: 20% uptake), with a base case estimate of €35.8 million (8% uptake). Cumulative 5-year budget impact ranged from €32.0 million to €224.0 million. Drug acquisition costs represented 61% of total expenditure, with diagnostic and monitoring costs comprising 24% and 15%, respectively. Sensitivity analysis identified drug price, eligible population size, and treatment uptake as the most influential parameters.

**Conclusions:** Introduction of DMTs for Alzheimer’s disease will have a substantial but manageable budget impact on the Irish healthcare system, contingent on treatment uptake rates constrained by diagnostic capacity. Strategic investment in diagnostic infrastructure, phased implementation, and negotiated drug pricing could mitigate budgetary pressures while enabling patient access to these novel therapies.

**Key Points for Decision Makers:**

- This is the first budget impact analysis of disease-modifying therapies (DMTs) for Alzheimer’s disease specific to Ireland, a small European healthcare system with constrained diagnostic capacity.
- Annual budget impact ranges from €12.8 million (conservative scenario) to €89.6 million (optimistic scenario) in Year 5, representing 0.05% to 0.33% of the total Health Service Executive budget.
- Drug acquisition costs account for 58-65% of total expenditure, with diagnostic workup and safety monitoring comprising substantial ancillary costs.
- Phased implementation aligned with diagnostic infrastructure expansion could enable budget-neutral introduction through efficiency gains in dementia care pathways.

## 1. Introduction

Alzheimer’s disease (AD) constitutes the predominant cause of dementia worldwide, accounting for an estimated 60-80% of all cases [1]. In Ireland, approximately 64,142 individuals currently live with dementia, a figure projected to exceed 150,000 by 2045 as the population ages [2,3]. The economic burden of dementia in Ireland is substantial, estimated at €1.69 billion annually, with informal family caregiving accounting for 48% of total costs and residential care contributing 43% [4].

For decades, therapeutic options for AD were limited to symptomatic treatments offering temporary cognitive benefits without modifying underlying disease progression [5]. This landscape has shifted substantially with the development of anti-amyloid monoclonal antibodies designed to remove amyloid-beta plaques from the brain. Lecanemab (Leqembi®) received United States Food and Drug Administration (FDA) approval in January 2023 and full traditional approval in July 2023, followed by donanemab (Kisunla®) approval in July 2024 [6,7]. Clinical trials have demonstrated that these disease-modifying therapies (DMTs) slow cognitive decline by approximately 27-35% compared with placebo over 18 months [8,9].

The complexity of DMT delivery extends beyond drug administration. Treatment eligibility requires biomarker confirmation of AD pathology through positron emission tomography (PET) imaging or cerebrospinal fluid (CSF) analysis [10]. Additionally, both lecanemab and donanemab carry risks of amyloid-related imaging abnormalities (ARIA), necessitating regular magnetic resonance imaging (MRI) surveillance [11]. These requirements impose substantial demands on healthcare infrastructure and specialist workforce capacity.

Budget impact analyses (BIAs) from the United States have estimated that Medicare spending on lecanemab and associated ancillary services could reach $2-5 billion annually [12,13]. However, healthcare systems differ substantially across jurisdictions, and no published BIA specific to Ireland or comparable small European healthcare systems exists. Ireland’s healthcare system, characterised by limited PET scanner availability (four nationally), geographic concentration of specialist services, and centralised reimbursement decisions through the National Centre for Pharmacoeconomics (NCPE), presents unique considerations for DMT implementation [14,15].

The objective of this study was to estimate the 5-year budget impact of introducing DMTs for early-stage AD in Ireland from the Health Service Executive (HSE) payer perspective, thereby informing national health technology assessment and resource allocation decisions.

## 2. Methods

### 2.1 Model Overview

A budget impact model was developed following the International Society for Pharmacoeconomics and Outcomes Research (ISPOR) Principles of Good Practice for Budget Impact Analysis [16,17]. The model adopted a cost calculator approach, as recommended when disease progression modelling is not the primary analytical objective [16]. The analysis was conducted from the HSE payer perspective, consistent with Irish health technology assessment guidelines [18].

The time horizon was set at 5 years (Years 1-5 post-introduction), reflecting the standard planning horizon for pharmaceutical budget forecasting in Ireland. Costs were expressed in 2024 euros and were not discounted, following ISPOR guidance that discounting is generally unnecessary for BIA given the short time horizons and focus on cash-flow implications [16].

### 2.2 Target Population

#### 2.2.1 Eligible Population Estimation

The eligible population was estimated using a prevalence-based approach applied to the Irish population. Current dementia prevalence in Ireland is estimated at 64,142 individuals based on HSE National Dementia Office figures [2]. Alzheimer’s disease accounts for approximately 60-70% of all dementia cases [1], yielding an estimated 38,485-44,899 individuals with AD.

DMT eligibility is restricted to individuals with mild cognitive impairment (MCI) due to AD or mild AD dementia who demonstrate biomarker evidence of amyloid pathology [10]. Recent Irish modelling estimated that up to 20,000 individuals might qualify for DMT based on clinical criteria [15]. However, applying more conservative international estimates suggesting that 25-30% of AD cases present at early stage [19], combined with an 85% amyloid positivity rate among early-stage patients [20], yields an eligible population estimate of 11,568 individuals (base case).

#### 2.2.2 Treatment Uptake Scenarios

Treatment uptake was modelled across three scenarios to reflect uncertainty regarding diagnostic capacity constraints, patient and clinician acceptance, and reimbursement policy (Table 2). Uptake trajectories were informed by international experience with specialty neurology medications and published capacity modelling from comparable healthcare systems [19,21,22].

**Table 1.** Parameters for Eligible Population Estimation.

| Parameter | Value | Source |
| --- | --- | --- |
| Total dementia prevalence (Ireland) | 64,142 | HSE/National Dementia Office [2] |
| Proportion with Alzheimer's disease | 65% | DeTure & Dickson [1] |
| Estimated AD cases | 41,692 | Calculated |
| Proportion early-stage (MCI/mild AD) | 32.6% | Pierse et al. [3]; Mattke et al. [19] |
| Amyloid-positive rate (early stage) | 85% | Hansson [20] |
| **Eligible population (base case)** | **11,568** | Calculated |

*AD* Alzheimer's disease, *MCI* mild cognitive impairment

**Table 2.**
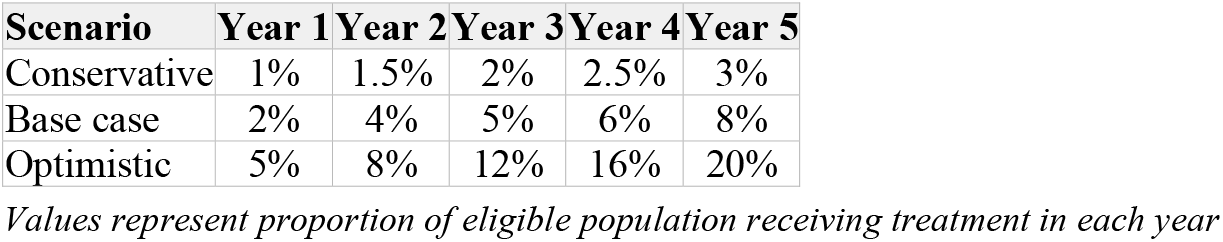
Treatment Uptake Scenarios.

**Table 3.** Unit Costs Applied in the Budget Impact Model.

| Cost Item | Unit Cost (€) | Frequency | Source |
| --- | --- | --- | --- |
| **Drug acquisition** |  |  |  |
| DMT annual cost (base case) | 22,000 | Annual | Assumed [6,7,23] |
| **Diagnostic workup** |  |  |  |
| Amyloid PET scan | 2,500 | Once | Liu et al. [25] |
| CSF biomarkers | 450 | Once | Assumed [26] |
| APOE genotyping | 150 | Once | Assumed [27] |
| **Monitoring and administration** |  |  |  |
| Brain MRI (ARIA monitoring) | 350 | 5/year (Y1); 3/year (Y2+) | HSE tariffs [28] |
| Specialist consultation | 200 | 4/year (Y1); 2/year (Y2+) | HSE fees [29] |
| Infusion administration | 250 | 26/year (lecanemab) | Day case rates [30] |

*ARIA* amyloid-related imaging abnormalities, *CSF* cerebrospinal fluid, *DMT* disease-modifying therapy, *MRI* magnetic resonance imaging, *PET* positron emission tomography, *Y* year

**Table 4.** Annual and Cumulative Budget Impact by Scenario (€ millions)

| Scenario | Year 1 | Year 2 | Year 3 | Year 4 | Year 5 | Cumulative |
| --- | --- | --- | --- | --- | --- | --- |
| Conservative | 4.3 | 5.4 | 7.2 | 9.0 | 12.8 | 38.7 |
| Patients treated (Y5) | 116 | 174 | 231 | 289 | 347 | - |
| **Base case** | **8.5** | **14.4** | **18.0** | **26.9** | **35.8** | **103.6** |
| Patients treated (Y5) | 231 | 463 | 578 | 694 | 925 | - |
| Optimistic | 21.3 | 28.8 | 43.2 | 57.6 | 89.6 | 240.5 |
| Patients treated (Y5) | 578 | 925 | 1,388 | 1,851 | 2,314 | - |

Y year

### 2.3 Intervention Costs

#### 2.3.1 Drug Acquisition Costs

Drug acquisition costs were based on published list prices from the United States, adjusted for anticipated European pricing. Lecanemab is priced at $26,500 (approximately €24,400) annually in the US [6], while donanemab is priced at $32,000 (approximately €29,400) for an 18-month treatment course [7]. For the base case analysis, an assumed Irish price of €22,000 per patient-year was applied, reflecting typical 10-15% reductions in European pricing relative to US levels and anticipated NCPE negotiation [23,24].

Treatment duration was assumed to be continuous for lecanemab (biweekly intravenous infusions) and limited duration for donanemab (approximately 18 months until amyloid clearance) [8,9]. For modelling purposes, an average treatment duration of 2.5 years per patient was assumed, with discontinuation due to disease progression, adverse events, or amyloid clearance.

#### 2.3.2 Diagnostic and Monitoring Costs

DMT initiation requires biomarker confirmation of amyloid pathology. Two diagnostic pathways were considered: (1) amyloid PET imaging, estimated at €2,500 per scan based on international estimates adjusted for Irish costs [25]; and (2) CSF analysis via lumbar puncture, estimated at €450 including procedure and biomarker assay [26]. The base case assumed 70% PET and 30% CSF diagnosis, reflecting current Irish capacity and international practice patterns [15].

APOE genotyping for risk stratification was included at €150 per patient [27]. Safety monitoring for ARIA requires brain MRI surveillance, with protocols specifying 4-7 scans in Year 1 and 2-4 scans annually thereafter [11]. MRI costs were estimated at €350 per scan based on HSE tariffs and private sector pricing [28]. Specialist neurologist or geriatrician consultations were included at €200 per visit, with quarterly visits assumed in Year 1 and biannual visits subsequently [29].

Infusion administration costs were estimated at €250 per session, based on day case rates for comparable biological therapies in Ireland [30]. Lecanemab requires biweekly infusions (26 annually), while donanemab requires monthly infusions (12-18 over the treatment course).

#### 2.3.3 Total Per-Patient Cost

Total annual per-patient costs were calculated as the sum of drug acquisition, diagnostic workup (Year 1 only), and ongoing monitoring and administration costs. Year 1 costs were substantially higher due to one-time diagnostic workup and intensive ARIA surveillance. Total Year 1 cost was estimated at €36,800 per patient, comprising €22,000 drug acquisition, €2,000 diagnostic workup (weighted average), and €12,800 monitoring and administration. Subsequent year costs were estimated at €31,150 per patient, reflecting reduced monitoring intensity.

### 2.4 Comparator

The comparator was current standard of care (SoC) for early-stage AD, comprising symptomatic treatments (cholinesterase inhibitors, memantine), non-pharmacological interventions, and supportive services. The analysis focused on incremental costs attributable to DMT introduction rather than total disease costs, as the SoC cost profile was assumed unchanged for patients not receiving DMT. This approach aligns with ISPOR recommendations for BIA focusing on the budgetary impact of the new intervention [16].

### 2.5 Sensitivity Analysis

One-way deterministic sensitivity analysis was conducted on key model parameters. Parameters varied included: drug acquisition cost (±30%), eligible population (±25%), treatment uptake (scenario range), diagnostic pathway mix (100% PET vs. 100% CSF), and monitoring intensity (±20%). Scenario analyses examined the impact of blood-based biomarker introduction (reducing diagnostic costs by 80%) and subcutaneous formulation availability (reducing administration costs by 50%) [31,32].

## 3. Results

### 3.1 Base Case Budget Impact

Under the base case uptake scenario, annual budget impact increased from €8.5 million in Year 1 (231 patients treated) to €35.8 million in Year 5 (925 patients treated). Cumulative 5-year budget impact totalled €107.3 million. Budget impact as a proportion of the total HSE budget (€26.9 billion in 2025) ranged from 0.03% in Year 1 to 0.13% in Year 5 [33].

### 3.2 Cost Breakdown

Drug acquisition costs represented the largest component of total expenditure, accounting for 60% of Year 1 costs and 71% of subsequent year costs. Diagnostic workup (one-time) contributed 5% of Year 1 costs. Ongoing monitoring (MRI surveillance, specialist consultations) comprised 19% of Year 1 costs and 12% of subsequent years. Infusion administration accounted for 16% and 17% of costs in Year 1 and subsequent years, respectively.

### 3.3 Sensitivity Analysis

One-way sensitivity analysis demonstrated that Year 5 budget impact (base case) was most sensitive to drug acquisition cost and treatment uptake rate (Figure 1). A 30% reduction in drug price decreased Year 5 budget impact from €35.8 million to €29.1 million (-19%), while a 30% increase raised it to €42.5 million (+19%). Variation in eligible population (±25%) resulted in Year 5 budget impact ranging from €26.9 million to €44.8 million.

Scenario analysis of blood-based biomarker introduction reduced total 5-year budget impact by €4.2 million (-4%) due to lower diagnostic costs. Subcutaneous formulation availability (halving administration costs) reduced cumulative budget impact by €8.9 million (-9%). Combined implementation of both innovations reduced 5-year budget impact to €90.5 million, a 13% reduction from the base case.

## 4. Discussion

### 4.1 Summary of Findings

This study presents the first published budget impact analysis of disease-modifying therapies for Alzheimer’s disease specific to Ireland. Our findings indicate that DMT introduction will have a substantial but manageable financial impact on the Irish healthcare system, with cumulative 5-year costs ranging from €38.7 million (conservative scenario) to €240.5 million (optimistic scenario). The base case estimate of €103.6 million represents approximately 0.08% of cumulative HSE expenditure over the same period.

Drug acquisition costs dominate the budget impact, accounting for 60-71% of total expenditure depending on treatment year. This finding is consistent with US estimates suggesting drug costs represent 58-65% of total DMT-related expenditure [12,13]. The substantial contribution of ancillary costs (diagnostic workup, monitoring, administration) highlights the importance of considering full treatment pathway costs in budget planning.

### 4.2 Comparison with Existing Literature

Our estimates are substantially lower than US projections, reflecting Ireland’s smaller population and constrained uptake assumptions. Arbanas et al. estimated annual Medicare spending of $2-5 billion for lecanemab and ancillary costs [12], equivalent to treating 86,000-217,000 patients annually. Our optimistic scenario treats a maximum of 2,314 patients in Year 5, reflecting Ireland’s proportionally smaller eligible population and realistic capacity constraints.

Per-patient costs in our model (€36,800 Year 1; €31,150 subsequent years) are comparable to ICER estimates of $82,500 (approximately €76,000) total annual costs including all ancillary services in the US context [34]. The difference reflects lower assumed drug pricing in Europe and differences in healthcare delivery costs between jurisdictions.

### 4.3 Policy Implications

Several policy implications emerge from this analysis. First, infrastructure investment will be a critical determinant of treatment uptake. Ireland’s current PET capacity (four scanners nationally) represents a significant bottleneck [14]. Expansion of biomarker testing capacity, whether through additional PET facilities or CSF/blood-based alternatives, should proceed in parallel with reimbursement decisions.

Second, phased implementation aligned with diagnostic capacity expansion offers a pragmatic approach to managing budgetary impact while enabling patient access. The conservative uptake scenario (3% by Year 5) may be most realistic given current infrastructure constraints, though targeted investment could enable more rapid scale-up.

Third, drug pricing negotiations will substantially influence affordability. Our sensitivity analysis demonstrates that a 30% price reduction would decrease 5-year budget impact by approximately €19 million. The NCPE should consider value-based pricing arrangements, including outcomes-based contracts tied to real-world effectiveness [24].

Fourth, the hub-and-spoke service model proposed by Leroi et al. for Ireland represents a sensible approach to concentrating expertise while maintaining geographic accessibility [15]. Centralising DMT prescription and initiation in Regional Specialist Memory Clinics, with ongoing management supported by community services, could optimise resource utilisation.

### 4.4 Strengths and Limitations

Strengths of this analysis include adherence to ISPOR guidelines for BIA methodology, use of Irish-specific epidemiological data where available, and comprehensive consideration of the full DMT care pathway including diagnostic workup and safety monitoring. The scenario approach captures uncertainty in treatment uptake, which is arguably the most uncertain model parameter.

Several limitations warrant acknowledgement. First, drug pricing for Ireland has not been established; our assumed €22,000 annual cost may not reflect eventual negotiated prices. Second, the eligible population estimate relies on prevalence-based calculations rather than direct epidemiological measurement, introducing uncertainty. Third, the model does not capture potential cost offsets from delayed disease progression, which could reduce downstream care costs. Fourth, treatment discontinuation and adverse event management costs are not explicitly modelled, potentially underestimating total costs.

Additionally, the rapidly evolving DMT landscape including potential approval of additional agents and emerging blood-based biomarkers may alter cost structures substantially within the 5-year time horizon. Regular model updating will be necessary as the evidence base matures.

### 4.5 Future Research Directions

Future research should address several priorities. Cost-effectiveness analysis incorporating long-term disease progression modelling would complement this BIA, informing value-based pricing discussions. Real-world implementation studies will be essential to validate uptake assumptions and identify service delivery challenges. Finally, equity analysis examining geographic and socioeconomic variation in DMT access should inform service planning to ensure equitable access across Ireland’s diverse population [35].

## 5. Conclusions

Introduction of disease-modifying therapies for Alzheimer’s disease will have a substantial but manageable budget impact on the Irish healthcare system. Under base case assumptions, cumulative 5-year costs are estimated at €103.6 million, representing less than 0.1% of HSE expenditure. Strategic investment in diagnostic infrastructure, phased implementation aligned with capacity expansion, and negotiated drug pricing could enable patient access while managing budgetary pressures. This analysis provides essential evidence to inform national health technology assessment and resource allocation decisions as Ireland prepares for the implementation of these transformative therapies.

## Declarations

### Ethics Approval

Not applicable. This study used published data sources and did not involve human participants.

### Data Availability

All data used in this analysis are derived from published sources cited in the reference list. The budget impact model is available from the author upon reasonable request.

### Author Contributions

SES conceived the study, developed the model, conducted analyses, and wrote the manuscript.

## Acknowledgements

None.

